# NSAIDS-XDR-TB clinical trial: A randomized pilot study to estimate the potential efficacy and safety of using adjunctive ibuprofen for the treatment of pre-XDR and XDR tuberculosis

**DOI:** 10.1101/2025.05.29.25328553

**Authors:** Kaori L. Fonseca, Judith Farrés, Ketevan Barbakadze, Iza Jikia, Malkhaz Tsotskhalashvili, Juan M. García-Illarramendi, Lilibeth Arias, Kennedy Otwombe, Chiara Sopegno, Albert Despuig, Neil Martinson, Nadiia Buhiichyk, Tamta Korinteli, Zaza Avaliani, Nestani Tukvadze, Sergo Vashakidze, Cristina Vilaplana

## Abstract

**Background:** Drug-resistant tuberculosis (TB) remains a formidable global health challenge. Host-directed therapies (HDTs) offer the potential to mitigate tissue damage, reduce treatment duration, and improve clinical outcomes by modulating immune responses. We assessed the safety and potential efficacy of adjunctive ibuprofen—an inexpensive, well-tolerated non-steroidal anti-inflammatory drug—in patients with pre-extensively drug-resistant (pre-XDR) and extensively drug-resistant (XDR) TB.

**Methods:** In this prospective, open-label, randomized pilot study (NCT02781909) conducted in Georgia, 28 adults with bacteriologically confirmed pulmonary pre-XDR or XDR-TB were randomized 1:1 to receive either standard-of-care (SoC) TB treatment alone (n=14) or SoC plus 400 mg ibuprofen daily during the first 2 months (n=14). Participants were followed for 6 months. The primary endpoints were early sputum culture conversion and radiological improvement. Secondary endpoints included WHO-defined final treatment outcomes, safety, health-related quality of life (HQoL), and changes in inflammatory markers.

**Findings:** By week 2, culture negativity was achieved in 27% of control participants versus 9% in the ibuprofen group (risk difference 18%, 95% CI −13 to 50). The median time to culture conversion was 4 months in both groups. At month 2, favourable X-ray evolution was observed in 64% of controls compared with 54% of the ibuprofen group (risk difference 9%, 95% CI −32 to 50), with 90% of participants in each group showing improvement by month 6. Final treatment outcomes were comparable (≈71% cured) and the incidence of safety-related events did not differ significantly. Notably, the ibuprofen group exhibited greater proportional reductions in inflammatory markers—including a statistically significant decrease in the monocyte-to-lymphocyte ratio at months 2 and 5, along with reductions in interferon gamma levels and the enrichment score for Thompson_FAIL_13 gene signature at month 6.

**Interpretation:** Although adjunctive ibuprofen did not improve primary microbiological or radiological endpoints, its excellent safety profile and significant anti-inflammatory effects support its potential role as an immune-modulating adjunct in the treatment of drug-resistant TB. These findings warrant further investigation in larger studies to optimize dosing and evaluate clinical benefits.

**Funding:** Catalan Government (2021 SGR 00920), Spanish Government-FEDER Funds (CPII18/00031, PI20/01424 and CB06/06/0031, and European Union’s Horizon 2020 research and innovation program under grant agreement No. 847762 (SMA-TB project).

**Research in context:** *Evidence before this study:* Drug-resistant tuberculosis (TB) remains a major global health challenge. Host-directed therapies (HDTs) have emerged as a means to shorten treatment, improve outcomes and limit tissue damage from excessive inflammation. In preclinical models of active TB, non-steroidal anti-inflammatory drugs (NSAIDs) enhanced bacterial control and reduced lung pathology. Clinical data on NSAIDs as HDTs are scarce, confined to small studies in tuberculous meningitis and TB patients with diabetes, which reported encouraging but preliminary benefits.

*Added value of this study:* This is the first randomized trial of adjunctive ibuprofen in patients with pre-XDR and XDR-TB. Whilst ibuprofen did not confer microbiological or clinical benefit at two and six months, it was safe, well tolerated and significantly lowered systemic inflammatory markers. Baseline imbalances in disease severity and high inter-patient variability likely obscured any clinical effect despite reduced inflammation.

*Implications of all the available evidence:* Our findings demonstrate that ibuprofen is a safe adjunct in drug-resistant TB and can mitigate harmful inflammation. Future work should explore optimised dosing regimens, alternative HDT candidates and more sensitive or tailored endpoints—potentially focusing on patient subgroups most likely to benefit. Such refinements will be essential to reveal the true clinical value of HDTs across diverse TB populations.

## Introduction

Tuberculosis (TB) disease is still claiming millions of lives annually, while imposing economic and social burdens, being one of the major problems for health systems worldwide. In 2024, nearly half a million TB cases emerged as multi-drug resistant (MDR) or extensively drug-resistant (XDR), with concerningly low success rates in treatment.^1^ Drug resistant (DR)-TB is a major public health challenge globally. Countries of eastern Europe and central Asia are home to 24% of the global cases of MDR-TB and 47% of the pre-extensively DR-TB cases (pre-XDR-TB).^2^ This poses a problem for European health systems, which suffer from the increased complications associated with these forms of the disease. Treatment for MDR/XDR-TB is prolonged, and is associated with poor outcomes and high mortality rates.^3^ Despite substantial efforts, achievements, and recent guidelines introducing new regimens, novel approaches to treat MDR-TB, reduce morbidity and mortality, shorten treatment duration, and prevent the emergence of new drug resistances, are still urgently needed.

Host immune responses and cellular processes are key to disease progression, making them valuable targets for developing new therapies that hinder pathogen survival and pathogenesis. Host-directed therapies (HDT) have emerged as a promising alternative to enhance treatment outcomes by modulating the immune system, tackling tissue damage and hyperinflammation, reducing treatment duration, and minimising drug-resistance risk.^4,5^ As hyperinflammation causes lung damage associated with worse outcomes and the presence of sequelae, the use of repurposed anti-inflammatory drugs as adjuncts to standard treatment is a promising option.^6^

Several drugs have been proposed as HDTs, based on *in vitro* studies and/or theoretical rationale. However, few have been tested in clinical trials and some—such as thalidomide, etanercept, and interferon gamma (IFNG)—are associated with severe adverse effects.^7^ Despite the urgency of the TB epidemic and its high mortality rate, low-risk HDT options remain largely unexplored in clinical trials.

Ibuprofen is a non-steroidal anti-inflammatory drug (NSAID) and a non-selective cyclooxygenase inhibitor (PGH1, PGH2, commonly known as COX-1 and COX2) that limits the biosynthesis of prostaglandins. It has been widely used for decades to relieve pain and fever, as well as to treat inflammatory diseases. Ibuprofen has a well-established safety profile and is highly affordable.^8^

Pre-clinical studies with over-the-counter NSAIDs have shown beneficial effects in a murine model of active TB.^9^ Ibuprofen has previously been shown to reduce lung pathology and mycobacterial burden in a highly susceptible mouse model of TB increasing its survival. In the absence of TB treatment, ibuprofen markedly and significantly increased survival and decreased both the bacillary load and granuloma in mice treated with ibuprofen.^10^ This effect is likely due to NSAIDs reducing proinflammatory mediators (TNF, IL-17, IL-6 and CXCL5) release rather than a direct toxic effect on mycobacteria.

Previous clinical studies on NSAIDs as HDT have also shown promising benefits, mainly in individuals suffering from tuberculous meningitis, though ibuprofen had never been tested before. ^11–13^ In this context, we hypothesized that adding ibuprofen to standard-of-care (SoC) TB treatment could provide additional benefits. The NSAIDS-XDR-TB pilot study aimed to assess the potential efficacy and safety of adjunctive ibuprofen in the treatment of pre-XDR and XDR-TB.

## Material and Methods

### Study design and participants

NSAIDS-XDRTB was designed as a phase IIA prospective, interventional, randomized (1:1), controlled, open-label, pilot trial to estimate the potential efficacy and safety of using adjunctive ibuprofen for the treatment of pre-XDR and XDR-TB (ClinicalTrials.gov NCT02781909). The trial was conducted in the National Center for TB and Lung Diseases (NCTLD – Georgia). Ethics approval for this study was granted by the Ethics review Committee of NCTLD-Georgia.

Eligible participants included males and females aged 16 or older with bacteriologically confirmed pulmonary pre-XDR or XDR-TB, with or without extrapulmonary involvement. Patients were excluded if they had a history of TB, HIV-1 infection, chronic hepatitis B virus infection, diabetes, abnormal chemistry or haematology values, or if they had required corticosteroids or other prohibited medications in the past 28 days. Before enrolment, all participants provided written informed consent. Detailed inclusion and exclusion criteria are available in Supplementary Table 1.

### Sample size

As a pilot study, the following trial was designed to assess the magnitude and direction of possible responses and not powered to show smaller differences. The total n of 12 per treatment arm was defined based on feasibility, gains in the precision about the mean and variance, and regulatory considerations, according to the literature on pilot studies.^14,15^

### Randomization and masking

At diagnosis, participants were enrolled and randomly assigned to one of the experimental groups in a 1:1 ratio. The allocation sequence was randomly generated by a computer-generated randomisation list. No placebo interventions were administered, so randomization was open (unblinded) to both participants and medical trial investigators.

### Procedures and interventions

The second-line treatment regimens were personalized and tailored either using optimized treatment based on molecular drug sensitivity testing (Line-Probe assays) or classical phenotypic drug susceptibility testing, following pre-XDR and XDR-TB guidelines (Supplementary Table 2). Participants were categorized as pre-XDR-TB if they were resistant to rifampicin and fluoroquinolones, and as XDR-TB if they were resistant to rifampicin, fluoroquinolones, and either bedaquiline or linezolid. They were randomly assigned to receive standard TB treatment (n=14) or standard TB treatment plus 400mg of orally administered ibuprofen daily for the first 2 months of therapy (n=14). The study medication was provided by the Pharmacy Department of the NCTLD and administered as per the clinical routine. Participants were followed for up to 6 months through standard visits and sputum examinations, as per clinical routine (Supplementary Table 3).

### Study Outcomes

The primary efficacy outcome was to determine the proportion of pilot study participants showing clinical or microbiological benefits from the new regimen relative to controls. These were measured by evaluating:

- percentage of negative sputum culture at month 2 and 6 following inclusion and time to stable sputum culture conversion (SCC) up to month 6: (≥ two consecutive cultures negative for *M. tuberculosis*) including all time in follow-up to month 6 on TB treatment.
- changes detected by X-ray during follow-up: at baseline and every 3 months (as per routine).
- final treatment outcomes according to the WHO definitions including all time in follow-up to 6 months on TB treatment. Participants were considered cured and successfully treated when showing evidence of bacteriological clearance with no reversion and no evidence of failure. Participants whose treatment was interrupted for two or more consecutive months were considered lost to follow-up.^16^

The secondary outcomes were to compare the proportion of study participants:

- who tolerated the new regimen. Outcome measurements: incidence of safety-related events during the whole study period: clinical worsening of the disease, no sputum conversion (if AFS-), any worsening concerning vital parameters and routine blood work.
- showing differences in Health Quality of Life (HQoL). Outcome measurements: HQoL measures months 2 and 6, relative to baseline.
- showing differences in immune responses at months 2 and 6 compared to baseline. Outcome measurements: changes detected in immune responses at months 2 and 6. The immunological studies were assessed in all randomized patients.

### Demographic and Clinical Data collection

Once the informed consent was obtained, data from the study was recorded in a case report form (CRF) created *ad hoc*. The recruiting site used paper source documents to record the data and periodically entered it into an eCRF. Data collected included demographic information, medical history, and concomitant medications. Radiological efficacy was assessed using a scoring system that aggregates all X-ray findings (X-ray score) reported in the CRF. Parenchymal abnormalities, both primary and secondary, along with nodular and pleural abnormalities were considered (Supplementary Table 4). TB disease severity was evaluated at baseline using a score adapted from the Bandim TB score^17^ (Supplementary Table 5). The Saint George Respiratory Questionnaire (SGRQ) and Kessler-10 (K10) questionnaire were used to measure patients’ health and well-being and, psychological distress, through anxiety and depressive symptoms measurements, respectively.^18,19^

### Immunological response analysis

Composite inflammation indices were calculated using CRF data. These indices integrate independent white blood cell subsets, including the neutrophil-to-lymphocyte ratio (NLR), the monocyte-to-lymphocyte ratio (MLR), and the systemic immune-inflammation index (SII), which is computed as NLR multiplied by the platelet count.

Peripheral whole blood samples were collected at baseline, and at months 2 and 6 of treatment. Plasma was isolated from whole blood collected in sodium citrate CPT tubes (BD Vacutainer® CPT™). Whole blood collected in PAXgene® tubes (PreAnalytiX GmbH, Hombrechtikon, Switzerland) were used for the RNA-sequencing analysis. Samples were stored at −80° until analysis.

Levels of circulating pro- and anti-inflammatory cytokines in plasma were assessed by immunoassay. Levels of tumour necrosis factor (TNF), interferon gamma (IFNG) and interleukins (IL) IL-1B, IL-2, IL-4, IL-6, IL-8, IL-10, IL-12, and IL-17A were measured using the Millipore’s MILLIPLEX MAP human high sensitivity T cell magnetic Bead panel (cat#HSTMAG-28SK, EMD Millipore corporation). The inflammatory markers were evaluated in a total of 73 plasma samples from the 28 enrolled participants at baseline, months 2 and 6. Assays were performed according to the manufacturer’s instructions and plates read using a Luminex® 200 System.

Total RNA was extracted from 73 whole blood samples using the PAXgene Blood miRNA Kit (PreAnalitiX, Hombrechtikon, Switzerland) procedure, according to the manufacturer’s protocol. Total RNA quality was quantified using the Quant-iT Broad-Range dsDNA Assay Kit (Thermo Fisher Scientific), and RNA integrity was assessed using the Fragment Analyzer system HS RNA Kit (15NT) (Agilent). RNA-Seq libraries were prepared with the Illumina Stranded Total RNA Prep with Ribo-Zero Plus (Illumina), following the manufacturer’s recommendations, starting with 0·5 μg of total RNA as the input material. The final library was validated on an Agilent 2100 Bioanalyzer using the DNA 7500 assay. The libraries were sequenced on the NovaSeq 6000 (Illumina) with a read length of 2×101 bp following the manufacturer’s protocol for dual indexing. Image analysis, base calling, and quality scoring of the run were performed using the manufacturer’s software Real Time Analysis (RTA v3.4.4), followed by the generation of FASTQ sequence files.

### RNA-Seq data analysis

To identify differentially expressed genes (DEGs) between treatment groups across time points, we used the log-likelihood ratio test implemented in the DESeq2 R package (v1.38.3).^20^ A full model including treatment, time point, and their interaction was compared to a reduced model containing only treatment and time point. DEGs were defined using an adjusted p-value threshold of ≤ 0.05, corrected via the Benjamini-Hochberg method. This approach identified genes whose expression varied significantly between the ibuprofen and control groups over time.

Additionally, seven external predictive gene signatures related to TB treatment (Supplementary Table 6) were extracted from the TBSignatureProfiler R package.^21^ An enrichment score for each signature was calculated for each sample using single-sample gene set enrichment analysis (ssGSEA).

### Statistical analysis

Both primary and secondary analyses followed an intention to treat (ITT) approach, including all randomized patients. Continuous data were presented as medians and interquartile ranges (IQRs) and compared using the Mann-Whitney U test, with Benjamini-Hochberg correction for multiple testing. Categorical data were summarized by study group as counts and percentages of participants. The absolute risk difference was calculated with 95% confidence intervals (CIs) using the Newcombe method for differences in proportions.

The primary outcomes included the measurement of changes detected by X-ray score during follow-up, relative to baseline. We evaluated the changes by determining the ratios at month 3 (M3/BL) and 6 (M6/BL), participants showing a reduction in the X-ray, ratios < 1 were considered to have a favourable outcome. For the secondary outcomes, participants were classified as good progressors if their health-related quality of life (HQoL) scores—measured by the SGRQ and K10—fell within the normal range at both months 2 and 6 (Supplementary Tables 7 and 8). Specifically, this was defined as an SGRQ score of 7 or less and a K10 score below 20. For immune response, we calculated the ratios at months 2 and 6 relative to baseline, and compared the null hypothesis of no difference between them using the Mann-Whitney U test.

All statistical tests were two-sided, with a significance level of 5% (α = 0.05). Statistical analyses were performed using GraphPad Prism (version 10.0), R studio v.4.3.1 and Python v3.11.6.

### Role of the funding source

The funders of the study had no role in study design, data collection, data analysis, data interpretation, or writing of the report.

## Results

### Participant characteristics and clinical data on TB episode

A total of 28 participants diagnosed with a recurrent episode of TB were screened and assessed for eligibility between October 2016 and February 2018. All participants were enrolled and randomly assigned to either receive the SoC TB treatment (control group, n=14) or the SoC plus Ibuprofen (ibuprofen-treated group, n=14) regimens. The NSAIDS-XDR-TB flow diagram is showed in Figure 1.

**Figure 1 -.**
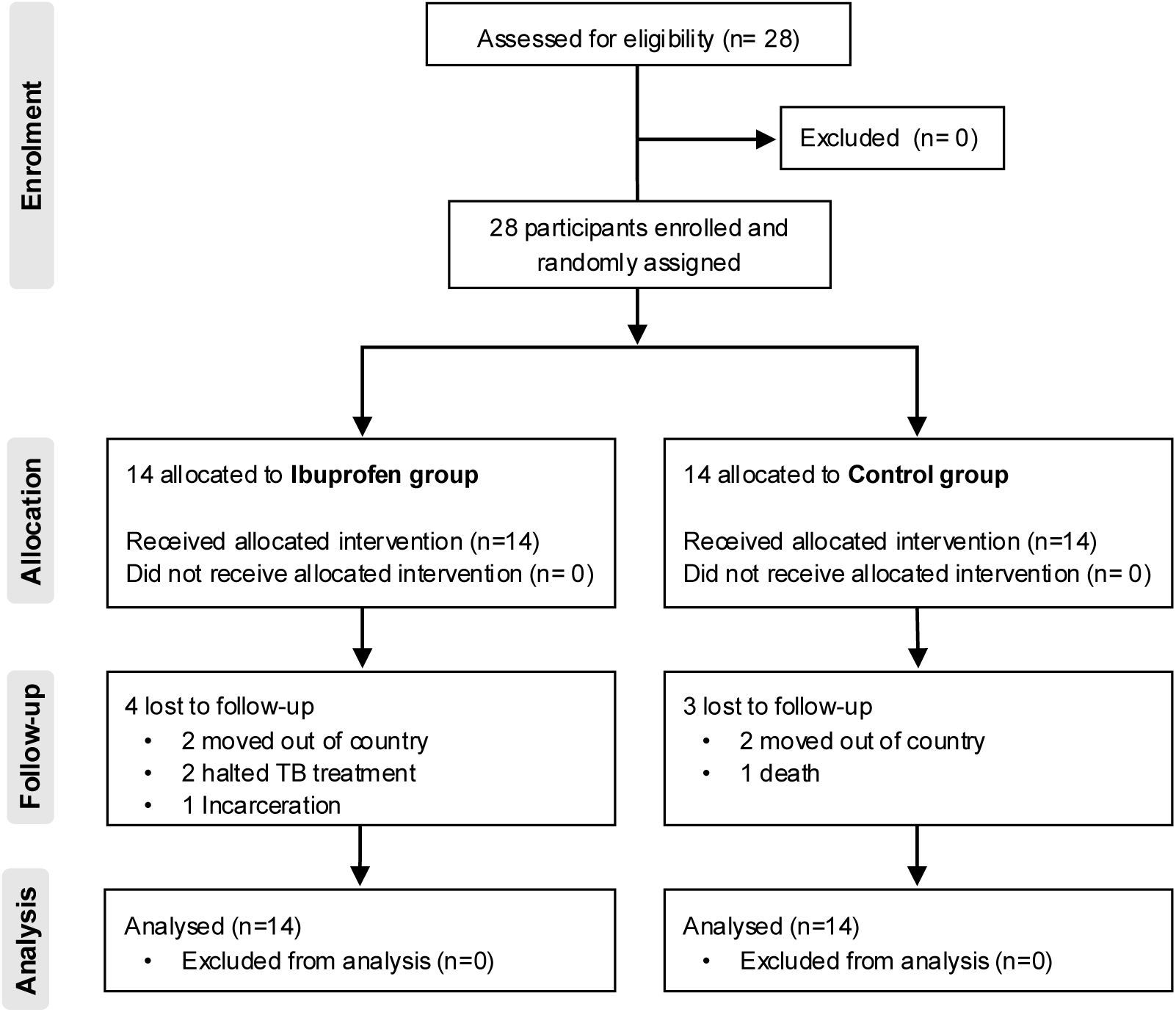
Study flow diagram. CONSORT flow diagram of the NSAIDS-XDR-TB pilot trial. Participants were assessed for eligibility, randomized, and allocated to either the control group, receiving the standard TB care, or the Ibu-treated group, receiving the standard care plus ibuprofen as an adjunctive therapy. All participant data was analysed.

The demographic and clinical baseline characteristics of the participants enrolled are presented in Table 1 and Supplementary Table 9. Participants’ median age was 44 years and 75% of the participants were male. The ibuprofen-treated group ended up with a higher proportion of female participants.

**Table 1.**
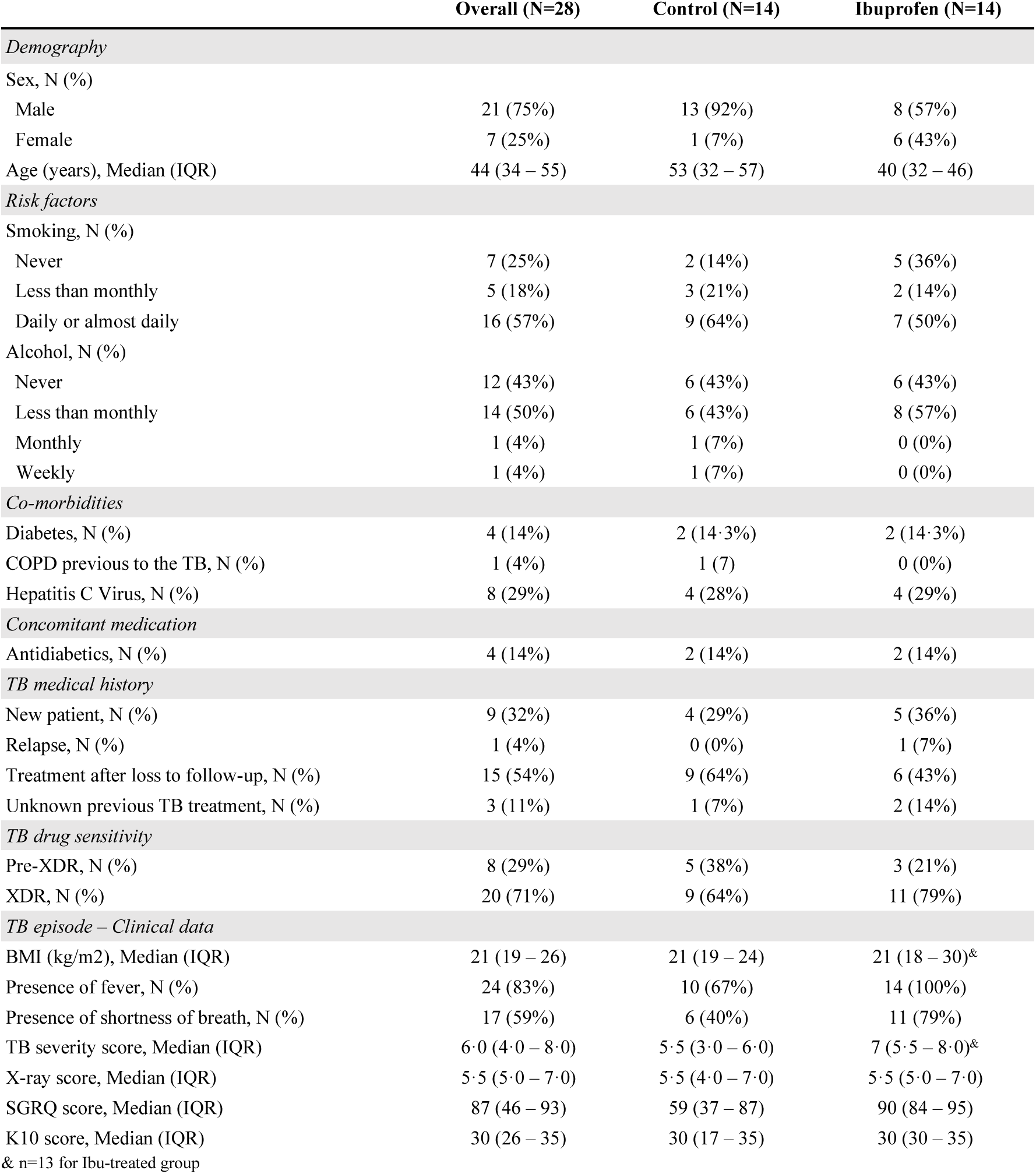
– Baseline characteristics by experimental group. COPD, Chronic obstructive pulmonary disease; BMI, body mass index; IQR, interquartile range; XDR, Extensively drug-resistant; TB, tuberculosis; SGRQ, Saint George Respiratory questionnaire; K10, Kessler Psychological Distress Scale.

Risk factors were related to smoking and alcohol consumption, being the proportions equivalent between experimental groups. The most frequently reported comorbidities, present in at least 14% of participants, were diabetes and hepatitis C virus co-infection. Considering the overall TB medical history of the participants, the majority (54%,15 out of 28) were treatment after lost to follow-up patients, and TB drug resistance profiles were comparable between the treatment groups. In the control group, five participants (38%, 5 out of 14) had pre-XDR-TB, and nine participants (64%, 9 out of 14) had XDR-TB. In the ibuprofen-treated group, three participants (21%, 3 out of 14) had pre-XDR-TB and eleven participants (79%, 11 out of 14) were XDR-TB.

The ibuprofen-treated group exhibited evidence of worse disease severity at baseline compared to the control group (Table 1). Specifically, participants in the ibuprofen-treated group had a baseline median TB severity score of 7 (IQR 5·5 – 8), whereas the control group had a score of 5·5 (IQR 3 – 6·25). The TB severity score is a clinical tool that quantifies TB disease severity based on various signs and symptoms, including fever and shortness of breath. Notably, these two symptoms also showed an imbalance between groups, with fever present in 67% (10 out of 14) of the control group vs. 100% of the ibuprofen-treated group and shortness of breath in 40% (6 out of 14) vs. 79% (11 out of 14), respectively.

Furthermore, the baseline severity differences were consistent with the SGRQ score, which was 59 (IQR 36 – 88·25) in the control group, compared to 90 (IQR 80 – 95·25) in the ibuprofen-treated group.

### Microbiological changes during follow-up

Microbiological efficacy was assessed by the proportion of participants achieving a stable negative sputum culture at month 2. No significant differences were found between groups: 9% (1 out of 11) of the ibuprofen-treated group versus 27% (3 out of 11) of the control group, with a risk difference (RD) of 18% and 95% CI (−13 to 50). By month 6, all participants in both groups had negative sputum cultures (Figure 2). The median time to sputum culture conversion was four months in both groups (Mann-Whitney test, p ≥ 0.5) (Figure 3). Median values are reported in Supplementary Table 10.

**Figure 2 -.**
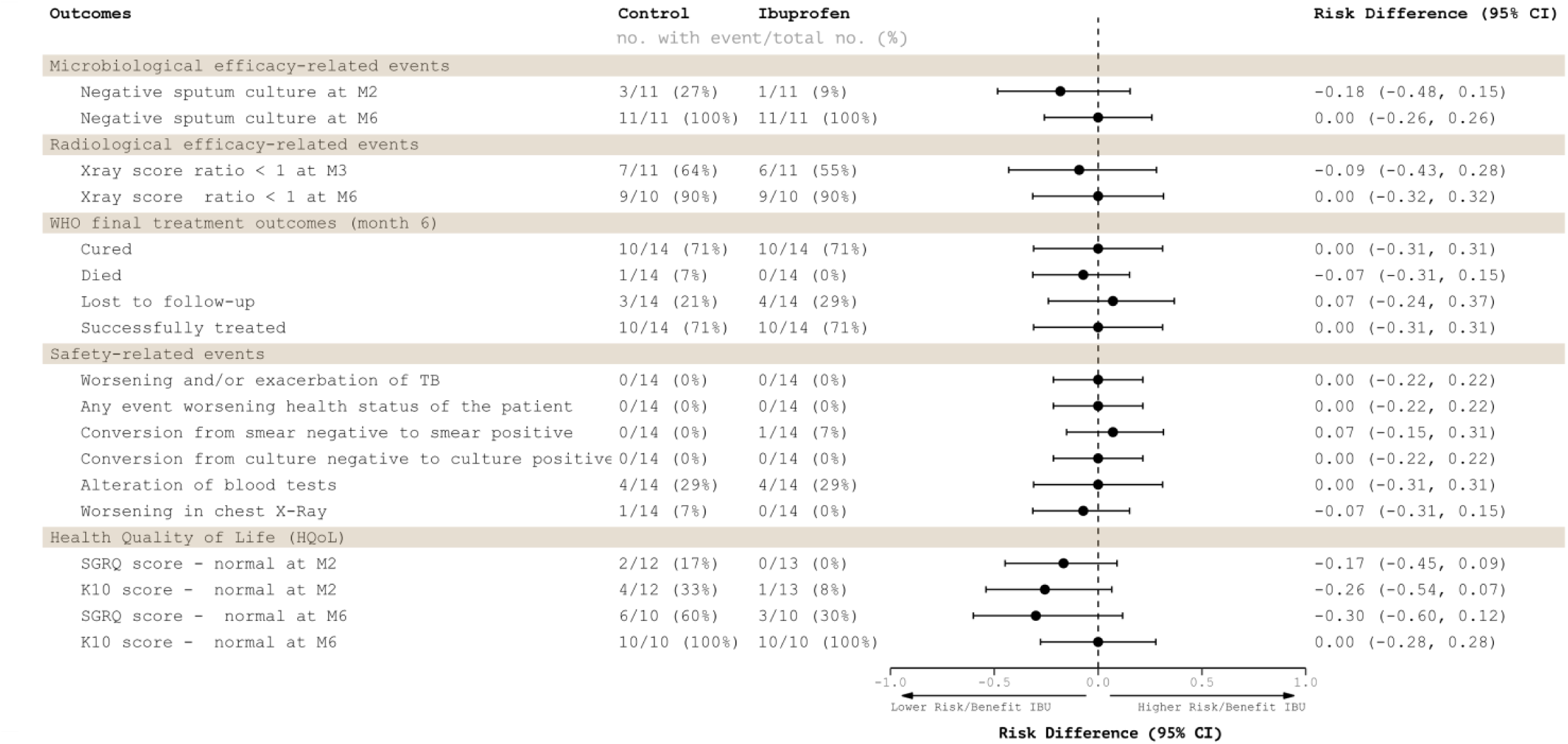
Primary and secondary outcomes risk difference. The 95% confidence intervals (CIs) for risk difference were calculated using the Newcombe method for differences in proportions. SGRQ, Saint George Respiratory questionnaire; K10, Kessler Psychological Distress Scale.

**Figure 3 -.**
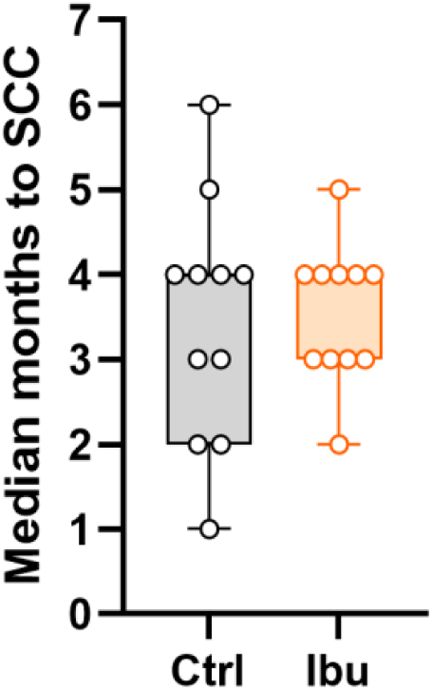
Primary outcome, time to stable sputum conversion. The median time to sputum culture conversion (SCC) in months for each treatment group is shown. Boxplots depict the median, interquartile ranges and minimum/maximum values. Each dot represents the ratio for a patient. Statistical analysis was performed using unpaired two-tailed Mann-Whitney test.

### Radiological changes during follow-up

Radiological efficacy was assessed using an X-ray score that accounted for key radiological signs, including primary and secondary parenchymal abnormalities, nodular lesions, and pleural abnormalities. No significant differences were observed between groups in the proportion of participants showing improvement over time. At month three, seven out of eleven participants in the control group and six out of eleven in the ibuprofen-treated group showed improvement, resulting in an RD of 9% (95% CI: −32 to 50). By month 6, all participants demonstrated some improvement compared to baseline, with no difference between groups (RD: 0%, 95% CI: −26 to 26) (Figure 2). Median values are reported in Supplementary Table 10 and ratios relative to baseline are presented in Supplementary Figure 1.

### Final treatment outcomes according to WHO definitions

When evaluating the proportion of participants on final treatment outcomes based on WHO definition, no significant differences were observed between experimental groups with a 71% achieved successful treatment. In the control group, 77% of participants were classified as cured (10 out of 14), 21% (3 out of 14) were lost to follow-up, and 8% (1 out of 13) died. In the ibuprofen-treated group, 71% (10 out of 14) of participants were cured, 29% (4 out of 14) were lost to follow-up, and no participant died (Figure 2).

### Safety-related events

Overall, the new regimen was well tolerated compared to the control, with all participants tolerating the regime. Also, the proportion of participants experiencing safety-related events (Figure 2) was similar, no significant differences were found between the treatment groups. During the study period, five out of 14 participants in each group experienced a safety-related event. One participant in the ibuprofen-treated group experienced a conversion from smear negative to smear positive but none converted from culture negative to culture positive. One participant in the control group showed worsening on the chest X-ray. The most frequent safety-related event was related to alterations of blood tests, affecting 28% (4 out of 14) of participants in each group.

### Changes in HQoL

HQoL was assessed by the SGRQ^18^ and K10 scores^19^. Participants who showed a score within the normal range (Supplementary Tables 7 and 8) at months 2 and 6 after treatment initiation were considered responders. No significant differences between study groups were observed at either time point for both scores (Figure 3). The proportion of participants showing a normal SGRQ score at month 2 were 17% (2 out of 12) in the control group, while none of the participants in the ibuprofen-treated group reached the normal score (0 out of 13) (Figure 2). At month 6, 60% (6 out of10) of the control group and 30% (3 out of 10) of the ibuprofen-treated group reached a normal score.

The proportion of participants not experiencing psychological distress at month 2, was 33% (4 out of 12) in the control group and 8% (1 out 13) in the ibuprofen-treated group. At month 6, all participants reached a normal score (Figure 2). Median values are reported in Supplementary Table 10 and ratios relative to baseline are presented in Supplementary Figure 1.

### Impact on the immune responses

The impact on the immune response was assessed by comparing the proportion of study participants showing changes in immune responses at months 2 and 6 relative to baseline. Three types of measures were used to evaluate the immune response: blood cell-derived inflammation index parameters (MLR, NLR, SII), plasma biomarkers related to immunological processes, and whole blood RNA transcripts.

Blood cell-derived inflammation indexes generally showed a decrease over time, with a greater decrease (lower median ratio) in the ibuprofen-treated group compared to the control group. This trend was statistically significant for MLR at month 2 (Mann-Whitney p-value: 0.01) and month five (Mann-Whitney p-value: 0.03) (Figure 4a, and Supplementary Figure 2).

**Figure 4 -.**
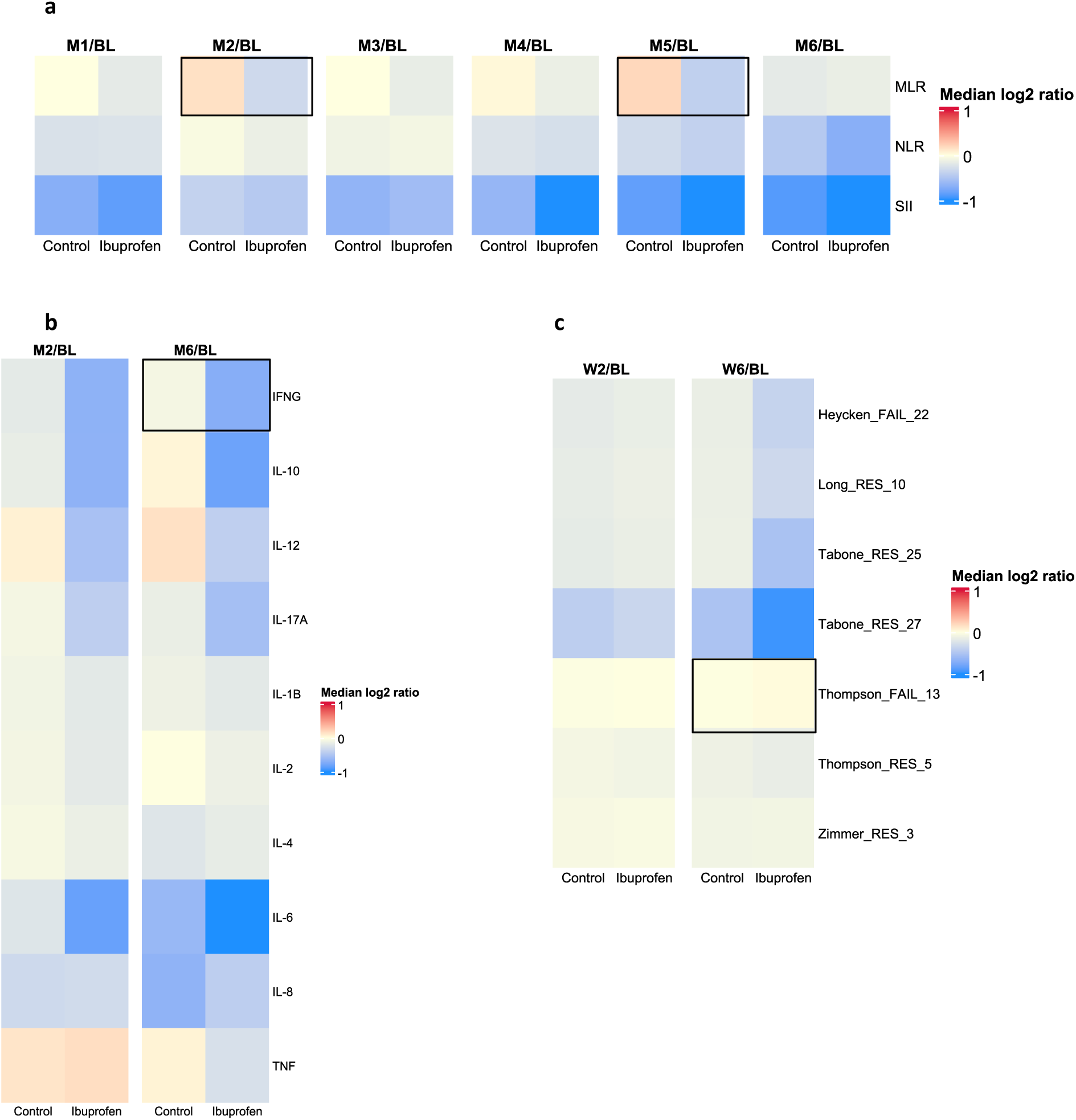
Immune response change along time. Heat maps depict the median change respect to baseline. Encapsulated cells denote p value<0·05 in unpaired two-tailed Mann-Whitney test **a.** Blood cell-derived inflammation index parameters change **b.** Plasma biomarkers change **c.** Enrichment score RNA based response signatures change.

Among the ten plasma biomarkers measured, IFNG, IL-10, IL-12, IL-17, and IL-6 followed a similar pattern, with the ibuprofen-treated group showing greater decreases at months 2 and 6 relative to baseline (Figure 4b, and Supplementary Figure 3). However, only IFNG at month 6 reached statistical significance (Mann-Whitney p-value: 0·045). IL-1B, IL-2, and IL-4 showed minimal decreases in both groups. IL-8 decreased more in the control group at month 6, while TNF decreased more in the ibuprofen-treated group at the same time, though none of these trends were statistically significant.

Analysis of RNA transcript levels over time did not reveal significant differences between the study groups. However, when comparing the gene expression profile to host blood RNA signatures that predict TB treatment outcomes, we observed a trend like the other immune measures, with the ibuprofen-treated group showing a greater decrease in the risk of treatment failure over time.

We selected seven published TB signature gene sets (Supplementary Table 6) that predict the risk of treatment failure. Using ssGSEA, we transformed each participant’s gene expression profile into scores for each selected TB signature and compared the changes in scores over time between the experimental groups. Higher gene signature scores correlate with more severe disease, except for the Thompson_FAIL_13 signature, where the correlation with disease severity is inversed, a higher score indicates less severity. At month 2, neither study group showed a decrease in the score for the analysed signatures, except for Tabone_RES_2, where both experimental groups showed a reduction in the score (Figure 4c, and Supplementary Figure 4). By month 6, differences between the experimental groups became more pronounced, with the ibuprofen-treated group showing a greater reduction in disease severity, reflected by a higher decrease in scores for all signatures, and an increase for Thompson_FAIL_13 where the less severity corresponds to a higher score. Due to small sample sizes and high variability, achieving significant results was challenging. The only comparison that reached statistical significance between groups was for Thompson_FAIL_13 at month 6 (Mann-Whitney p-value: 0.03).

## Discussion

HDTs have emerged as promising adjuncts to standard TB therapy, with the potential to reduce inflammation, enhance antibiotic efficacy, and improve patient outcomes. In this study, we present the first evaluation of the potential efficacy and safety of adjunctive ibuprofen in treating pre-XDR and XDR-TB cases. Our hypothesis was that adding ibuprofen to the SoC could provide additional benefits. Treatment efficacy and safety were assessed using WHO-endorsed tools— including chest X-rays, the SGRQ, and the K10 psychological distress score—as well as evaluations of immune responses.

In terms of clinical efficacy, our study did not demonstrate significant differences between patients treated with adjunctive ibuprofen and those receiving SoC alone. Similar percentages of participants in both treatment arms achieved negative sputum cultures at months 2 and 6, and the median time to stable SCC was four months in each group—a delay commonly observed in drug-resistant TB ^22^. Radiological assessments revealed no significant differences in chest X-ray evolution, despite the ibuprofen-treated group initially presented with more severe disease. Likewise, WHO-defined treatment outcomes indicated that 71% of participants in both arms were classified as cured or successfully treated, and no significant differences were found in HQoL outcomes.

Ibuprofen was well tolerated, with no significant differences in the incidence of safety-related events between groups. Ibuprofen is known for its strong anti-inflammatory properties and effective pain relief, with a relatively low risk of adverse effects, particularly in comparison to other NSAIDs^8^. The low dose used in this pilot study was chosen based on preclinical studies, the available safety literature, and potential interactions with other drugs in the SoC regimen. While this low dosage likely contributed to the excellent safety profile—given that NSAID tolerability is dose-dependent—it may also explain the lack of a significant increase in clinical efficacy. Moreover, the small sample size (28 individuals, with seven lost to follow-up) may have led to chance imbalances; notably, the ibuprofen-treated group had a higher proportion of female participants and a greater baseline severity of TB. Despite these imbalances, the comparable outcomes between groups at the end of treatment suggest that the addition of ibuprofen may have been beneficial.

Analysis of immune markers revealed a general trend toward greater reductions in inflammatory indices in the ibuprofen-treated group. The small sample size and natural variability in immune responses may have limited the detection of more robust statistical differences.

The decrease in MLR achieved in the ibuprofen-treated group was statistically significant at months 2 and 5. Notably, MLR, NLR, and SII have been explored as markers of disease progression in various diseases and also in TB.^23^ Elevated MLR values have been linked to increased TB severity ^24^, and have been used in clinical trials to evaluate HDTs.^25^ Additionally, a study assessing a PGH2 inhibitor in TB demonstrated reduced pro-inflammatory responses— evidenced by diminished phosphorylation potential and signal transduction in monocytes^26^ — supporting the potential relevance of our findings.

Plasma biomarker analysis also showed a trend toward greater reductions in inflammation markers in the ibuprofen-treated group, although only IFNG at month 6 reached statistical significance. Interestingly, both pro-inflammatory cytokines (e.g., IFNG) and anti-inflammatory cytokines (e.g., IL-10) followed a similar downward trajectory. Prostaglandins, particularly PGE2, play a central role in regulating the production of both pro-inflammatory and anti-inflammatory cytokines. By suppressing PGE2 synthesis through PTGS1 and PTGS2 inhibition, ibuprofen disrupts these feedback loops, leading to reduced levels of both cytokine types. In contrast, cytokines such as IL-1B, IL-2, and IL-4 remained largely unchanged, likely due to their regulation via alternative signalling pathways that are independent of prostaglandin-mediated mechanisms.^27^

Furthermore, we evaluated participants’ transcriptional profiles using seven established blood RNA signatures predictive of TB treatment failure. At month 6, the ibuprofen-treated group demonstrated a greater reduction in risk scores for most signatures compared to the control group, with the Thompson_FAIL_13 signature reaching statistical significance. The genes involved in this signature are enriched in neutrophil-associated transcripts and expressed higher in treatment failures at baseline, suggesting the presence of a state of enhanced inflammation prior to treatment in patients who will ultimately not be cured.^28^

Despite reductions in immune activation, these changes did not translate into improved microbiological or clinical outcomes. Several factors may explain this discrepancy. The degree of inflammation reduction achieved by ibuprofen might have been insufficient to significantly impact TB progression, possibly due to the low dose used or the specific NSAID selected—as other HDTs have been shown to contribute to inflammation resolution and at the same time to significantly improve sputum conversion rates and chest radiological outcomes.^25^

In addition to our study, several trials have evaluated adjunct anti-inflammatory treatments (aspirin, dexamethasone and prednisolone) in TB for tuberculous meningitis (NCT04145258, NCT03927313 and NCT03092817), for tuberculous pericarditis (NCT00810849) and pulmonary TB and type 2 diabetes.^13^ Moreover, two registered clinical trials on ClinicalTrials.gov have focused on NSAIDs for pulmonary TB: the TBCOX2 Trial (NCT02503839)^29^, which evaluated etoricoxib (a PTGS2 selective inhibitor), and the SMA-TB RCT (NCT04575519)^30^, which evaluated a higher dose of ibuprofen. Although results from the SMA-TB trial are not yet available, etoricoxib appears to be safe; however, the data did not support the use of adjunctive PTGS2 inhibitors, with the authors acknowledging the heterogeneity among TB patients and suggesting that cyclooxygenase inhibitors may benefit specific subgroups.^29^ Optimizing dosing or exploring alternative anti-inflammatory agents may be necessary to achieve a more pronounced clinical effect. Moreover, longer follow-up periods may be required to assess whether immune modulation confers sustained benefits in TB treatment and if any of these prevent TB relapse.

In conclusion, while our findings highlight ibuprofen’s potential as an immune-modulating therapy, further research is required to clarify its clinical relevance and optimize its therapeutic use in DR-TB. The limited sample size and single-site design of our pilot study constrain the generalizability of our findings. Future studies should explore alternative dosing schemes, additional HDT candidates, or combination approaches to enhance treatment efficacy. Given the empirical widespread clinical use of NSAIDs and the growing body of evidence supporting adjunctive anti-inflammatory therapy, more evidence and consensus on dosing, regimen, and patient selection is essential before these strategies can be incorporated into clinical guidelines.

## Author Contributions

CV conceptualised the study and acquired the funding. She developed the protocol with SV and NT and conduct project administration with LA and NT. KB, IJ, MT, NB, TK, ZA, NT and SV enrolled participants and oversaw clinical follow-up and data collection at their site. KO and NM carried out the primary statistical analysis. KLF, JF, JM G-I, CS, AD and NB performed the analysis of the immune responses. JF, JM G-I and CS performed the secondary outcomes statistical analysis. SV and CV supervised the project. KLF, JF and CV drafted the manuscript. CV was responsible for the decision to submit the manuscript. All authors contributed to data interpretation, critical review, and revision of the manuscript, and approved the decision to submit for publication. All authors provided written comments and feedback during manuscript development and were directly involved in the execution of the study.

## Declaration of interest

JMGI, CS, and JF were employees of Anaxomics Biotech S.L., Barcelona, during part of the study. CS and JF are now at IGTP, Badalona, and JMGI is at VHIR, Barcelona. CV declares receiving funding for the present study (2021 SGR 00920, CPII18/00031, PI20/01424, CB06/06/0031, and SMA-TB project (GA 847762), all paid to her institution. She is also a non-paid board member of 2 NPO: the foundation FUITB (http://www.uitb.cat/fuitb/) and ACTMON foundation (http://www.actmon.org/index.php), and the Secretary of TB & NTM group of the European Respiratory Society Assembly 10.

## Data sharing

To submit inquiries related to NSAIDS-XDR-TB clinical research, or to request access to individual participant data associated, please contact corresponding author.

RNA seq data has been submitted to the GEO database (accession number pending).

## Data Availability

To submit inquiries related to NSAIDS-XDR-TB clinical research, or to request access to individual participant data associated, please contact corresponding author.
RNA seq data has been submitted to the GEO database (accession number pending).

## Acknowledgments

We would like sincerely to thank the study participants who agreed to be part of the NSAIDS-XDR-TB study and the staff from the NCTLD who conducted the pilot clinical trial. We thank the CNAG sequencing platform for performing the RNA sequencing on the NovaSeq 6000 (Illumina) and for preparing the RNA-seq libraries. CNAG has the support of the Spanish Ministry of Science and Innovation through the Instituto de Salud Carlos III and the 2014–2020 Smart Growth Operating Program and cofinancing with the European Regional Development Fund (MINECO/FEDER, BIO2015-71792-P), and the support of the Generalitat de Catalunya through the Departament de Salut and Departament d’Empresa i Coneixement.

## Funding

This study received support from the Catalan Government (through 2021 SGR 00920), from the Spanish Government-FEDER Funds (through CPII18/00031, PI20/01424 and the CIBER Enfermedades Respiratorias (CB06/06/0031)), and through the European Union’s Horizon 2020 research and innovation program under grant agreement No. 847762 (SMA-TB project).

JMGI and CS acknowledge funding from the European Union’s Horizon 2020 research and innovation programme under the Marie Skłodowska-Curie grant agreement No 859962 and No 956148, respectively.

## Declaration of generative AI and AI-assisted technologies in the writing process

During the preparation of this work the authors used ChatGPT (OpenAI, ChatGPT, version 4) to improve language, readability and clarity of this manuscript. After using this tool, the authors reviewed and edited the content of the text and take full responsibility for the content of the publication.

## Notes

### Clinical Trial

NCT02781909

### Funding Statement

This study received support from:
-the Catalan Government (through 2021 SGR 00920)-the Spanish Government-FEDER Funds (through CPII18/00031, PI20/01424 and the CIBER Enfermedades Respiratorias (CB06/06/0031))
-the European Union Horizon 2020 research and innovation program under grant agreement No. 847762 (SMA-TB project).
JMGI and CS acknowledge funding from the European Union Horizon 2020 research and innovation programme under the Marie Skłodowska-Curie grant agreement No 859962 and No 956148, respectively.

### Author Declarations

Ethics approval for this study was granted by the Ethics review Committee of National Center for Tuberculosis Lung Diseases of Tbilisi,Georgia, where the trial was conducted.

